# Mendelian randomisation identifies alternative splicing of the FAS death receptor as a mediator of severe COVID-19

**DOI:** 10.1101/2021.04.01.21254789

**Authors:** Lucija Klaric, Jack S. Gisby, Artemis Papadaki, Marisa D. Muckian, Erin Macdonald-Dunlop, Jing Hua Zhao, Alex Tokolyi, Elodie Persyn, Erola Pairo-Castineira, Andrew P Morris, Anette Kalnapenkis, Anne Richmond, Arianna Landini, Åsa K. Hedman, Bram Prins, Daniela Zanetti, Eleanor Wheeler, Charles Kooperberg, Chen Yao, John R. Petrie, Jingyuan Fu, Lasse Folkersen, Mark Walker, Martin Magnusson, Niclas Eriksson, Niklas Mattsson-Carlgren, Paul R.H.J. Timmers, Shih-Jen Hwang, Stefan Enroth, Stefan Gustafsson, Urmo Vosa, Yan Chen, Agneta Siegbahn, Alexander Reiner, Åsa Johansson, Barbara Thorand, Bruna Gigante, Caroline Hayward, Christian Herder, Christian Gieger, Claudia Langenberg, Daniel Levy, Daria V. Zhernakova, J. Gustav Smith, Harry Campbell, Johan Sundstrom, John Danesh, Karl Michaëlsson, Karsten Suhre, Lars Lind, Lars Wallentin, Leonid Padyukov, Mikael Landén, Nicholas J. Wareham, Andreas Göteson, Oskar Hansson, Per Eriksson, Rona J. Strawbridge, Themistocles L. Assimes, Tonu Esko, Ulf Gyllensten, J. Kenneth Baillie, Dirk S. Paul, Peter K. Joshi, Adam S. Butterworth, Anders Mälarstig, Nicola Pirastu, James F. Wilson, James E. Peters

**Affiliations:** MRC Human Genetics Unit, Institute of Genetics and Cancer, University of Edinburgh, Western General Hospital, Crewe Road, Edinburgh, UK; Department of Immunology and Inflammation, Faculty of Medicine, Imperial College London, London, UK; Centre for Global Health Research, Usher Institute, University of Edinburgh, Teviot Place, Edinburgh, UK; British Heart Foundation Cardiovascular Epidemiology Unit, Department of Public Health and Primary Care, University of Cambridge, Cambridge, UK; Department of Human Genetics, Wellcome Sanger Institute, Hinxton, UK; Roslin Institute, University of Edinburgh, Easter Bush, Edinburgh, UK; Centre for Genetics and Genomics Versus Arthritis, Centre for Musculoskeletal Research, The University of Manchester, Manchester, UK; Institute of Genomics, University of Tartu, 51010, Estonia; Department of Medicine, Karolinska Institute, Stockholm, Sweden; Pfizer Worldwide Research, Development and Medical, Sweden; Department of Medicine, Stanford University School of Medicine, Stanford, CA, USA; MRC Epidemiology Unit, Institute of Metabolic Science, University of Cambridge School of Clinical Medicine, Cambridge, UK; Division of Public Health Sciences, Fred Hutchinson Cancer Research Center, Seattle, WA, USA; Population Sciences Branch, National Heart, Lung, and Blood Institute, National Institutes of Health, Bethesda, MD, USA; Framingham Heart Study, Framingham, MA, USA; Institute of Cardiovascular and Medical Sciences, University of Glasgow, Glasgow, UK; Department of Genetics, University of Groningen, University Medical Center Groningen, Groningen, the Netherlands; Department of Pediatrics, University of Groningen, University Medical Center Groningen, Groningen, the Netherlands; Danish National Genome Center, Copenhagen, Denmark; Faculty of Medical Sciences, Newcastle University, Newcastle upon Tyne, UK; Department of Clinical Sciences, Lund University, Malmö, Sweden; Wallenberg Center for Molecular Medicine, Lund University, Sweden; Hypertension in Africa Research Team (HART), North West University, Potchefstroom, South Africa; Uppsala Clinical Research Center (UCR), Uppsala University, Uppsala, Sweden; Clinical Memory Research Unit, Faculty of Medicine, Lund University, Lund, Sweden; Department of Neurology, Skåne University Hospital, Lund University, Lund, Sweden; Department of Immunology, Genetics and Pathology, Uppsala University, Sweden; Department of Medical Sciences, Uppsala University, Uppsala, Sweden; Department of Medical Epidemiology and Biostatistics, Karolinska Institutet, Stockholm, Sweden; Institute of Epidemiology, Helmholtz Zentrum München, German Research Center for Environmental Health, München-Neuherberg, Germany; German Center for Diabetes Research (DZD), München-Neuherberg, Germany; Division of Cardiovascular Medicine, Department of Medicine, Karolinska Institutet, Stockholm, Sweden; Institute for Clinical Diabetology, German Diabetes Center, Leibniz Center for Diabetes Research at Heinrich Heine University Düsseldorf, Düsseldorf, Germany; Division of Endocrinology and Diabetology, Medical Faculty, Heinrich Heine University Düsseldorf, Düsseldorf, Germany; Research Unit of Molecular Epidemiology, Helmholtz Zentrum München - German Research Center for Environmental Health, Neuherberg, Germany; Computational Medicine, Berlin Institute of Health (BIH) at Charité – Universitäts Medizin Berlin, Germany; Health Data Research UK, Wellcome Genome Campus and University of Cambridge, Cambridge, UK; Laboratory of Genomic Diversity, Center for Computer Technologies, ITMO University, St. Petersburg, Russia; Department of Cardiology, Clinical Sciences, Lund University; Skåne University Hospital, Lund, Sweden; Lund University Diabetes Center, Lund University, Lund, Sweden; The Wallenberg Laboratory/Department of Molecular and Clinical Medicine, Institute of Medicine, Gothenburg University; Department of Cardiology, Sahlgrenska University Hospital, Gothenburg, Sweden; The George Institute for Global Health, University of New South Wales, Sydney, Australia; Department of Surgical Sciences, Unit of Medical Epidemiology, Uppsala University, Uppsala, Sweden; Department of Physiology and Biophysics, Weill Cornell Medicine-Qatar, Doha, Qatar; Uppsala Clinical Research Center, Uppsala University, Uppsala, Sweden; Division of Rheumatology, Department of Medicine Solna, Karolinska Institutet, Sweden; Karolinska University Hospital, Stockholm, Sweden; Institute of Neuroscience and Physiology, University of Gothenburg, Gothenburg, Sweden; Memory Clinic, Skåne University Hospital, Malmö, Sweden; Institute of Health and Wellbeing, College of Medicine, Veterinary and Life Sciences, University of Glasgow, UK; Palo Alto VA Healthcare System, Palo Alto, CA, USA; Intensive Care Unit, Royal Infirmary of Edinburgh, 54 Little France Drive, Edinburgh, EH16 5SA, UK; British Heart Foundation Centre of Research Excellence, Addenbrookes Hospital, Cambridge, UK; National Institute for Health Research Blood and Transplant Research Unit in Donor Health and Genomics, University of Cambridge, Cambridge, United Kingdom

## Abstract

Severe COVID-19 is characterised by immunopathology and epithelial injury. Proteomic studies have identified circulating proteins that are biomarkers of severe COVID-19, but cannot distinguish correlation from causation. To address this, we performed Mendelian randomisation (MR) to identify proteins that mediate severe COVID-19. Using protein quantitative trait loci (pQTL) data from the SCALLOP consortium, involving meta-analysis of up to 26,494 individuals, and COVID-19 genome-wide association data from the Host Genetics Initiative, we performed MR for 157 COVID-19 severity protein biomarkers. We identified significant MR results for five proteins: FAS, TNFRSF10A, CCL2, EPHB4 and LGALS9. Further evaluation of these candidates using sensitivity analyses and colocalization testing provided strong evidence to implicate the apoptosis-associated cytokine receptor FAS as a causal mediator of severe COVID-19. This effect was specific to severe disease. Using RNA-seq data from 4,778 individuals, we demonstrate that the pQTL at the *FAS* locus results from genetically influenced alternate splicing causing skipping of exon 6. We show that the risk allele for very severe COVID-19 increases the proportion of transcripts lacking exon 6, and thereby increases soluble FAS. Soluble FAS acts as a decoy receptor for FAS-ligand, inhibiting apoptosis induced through membrane-bound FAS. In summary, we demonstrate a novel genetic mechanism that contributes to risk of severe of COVID-19, highlighting a pathway that may be a promising therapeutic target.

## Main

Severe COVID-19 is characterised by exaggerated inflammatory responses and immunopathology^1-4^. The two pharmacological treatments that have robustly demonstrated efficacy in reducing risk for severe COVID-19 in randomised clinical trials to date are glucocorticoids and interleukin 6 (IL-6) receptor blockade^5-7^. Treatments directed at the inflammatory response thus represent the most promising therapeutic strategy. A wide range of therapies directed at specific elements of the inflammatory response have been developed for autoimmune and inflammatory diseases^8,9^, and present potential repurposing opportunities for treatment of COVID-19. Profiling of plasma proteins in COVID-19 patients has revealed a signature of innate immune cell activation (including upregulation of IL-6, monocyte chemokines and neutrophil proteins) and epithelial/endothelial injury in severe disease^10,11^. A limitation of such observational studies, however, is that they cannot distinguish causal mediators of immunopathology from secondary downstream consequences of inflammation and/or tissue injury. Bridging this knowledge gap is critical for prioritising therapeutic targets and triaging medicines for clinical trials.

Making causal inference from human observational data is challenging due to confounding and reverse causation. Mendelian randomisation (MR) is an analytical approach that can circumvent these difficulties. MR enables causal inference by leveraging the random allocation of alleles at meiosis, which effectively provides a natural randomised trial^12,13^. MR tests whether there is a causal relationship between an exposure (e.g. a molecular trait) and an outcome (e.g. a clinical phenotype) using genetic variants as ‘instruments’. If a genetic variant associated with the exposure is also associated with the outcome, this provides evidence of a putatively causal relationship between the two. Using proteins as exposures in MR analyses has several advantages. First, proteins are gene products and as such are under greater genetic control than downstream phenotypes. Second, proteins are the targets of most drugs and so MR using proteins can identify and prioritise promising therapeutic targets. MR using proteins is now increasingly possible because of large genome-wide association studies (GWAS) that have identified genetic variants associated with levels of circulating proteins (protein quantitative trait loci, pQTL)^14-16^.

Here, we performed MR analysis to test whether proteins observationally associated with COVID-19 severity play a causal role in critical illness from COVID-19, using pQTL identified through a meta-analysis of up to 26,494 individuals. Our results implicate the cytokine receptor FAS as playing a putatively causal role in severe COVID-19. We demonstrated the robustness of this result using a range of sensitivity analyses and colocalisation analysis, and we replicated it using an independent pQTL dataset. The pQTL for FAS in the *FAS* gene region could not be explained by a corresponding expression quantitative trait locus (eQTL). We therefore examined mRNA splicing events in whole blood RNA-seq data from 4,778 individuals. This revealed that the pQTL for FAS in the *FAS* gene region is mediated by genetically influenced alternative splicing, resulting in skipping of exon 6 and affecting the ratio of soluble to membrane-bound FAS. We thus demonstrate a novel genetic mechanism contributing to risk of severe COVID-19 via a splice QTL (sQTL). We hypothesise that modulating the FAS pathway may therefore be a promising therapeutic strategy.

## Results

We first identified a list of proteins associated with COVID-19 clinical severity grading by examining two studies that performed broad proteomic profiling of COVID-19 patient plasma samples using the Olink proteomics platform^10,11^. We took the lists of severity-associated proteins from each study (Benjamini-Hochberg adjusted P <0.05) and intersected these to provide a high-confidence list of 157 severity-associated proteins (**Fig. 1**). To evaluate whether these proteins play a causal role in severe COVID-19, we performed two-sample Mendelian randomisation analysis.

**Figure 1:**
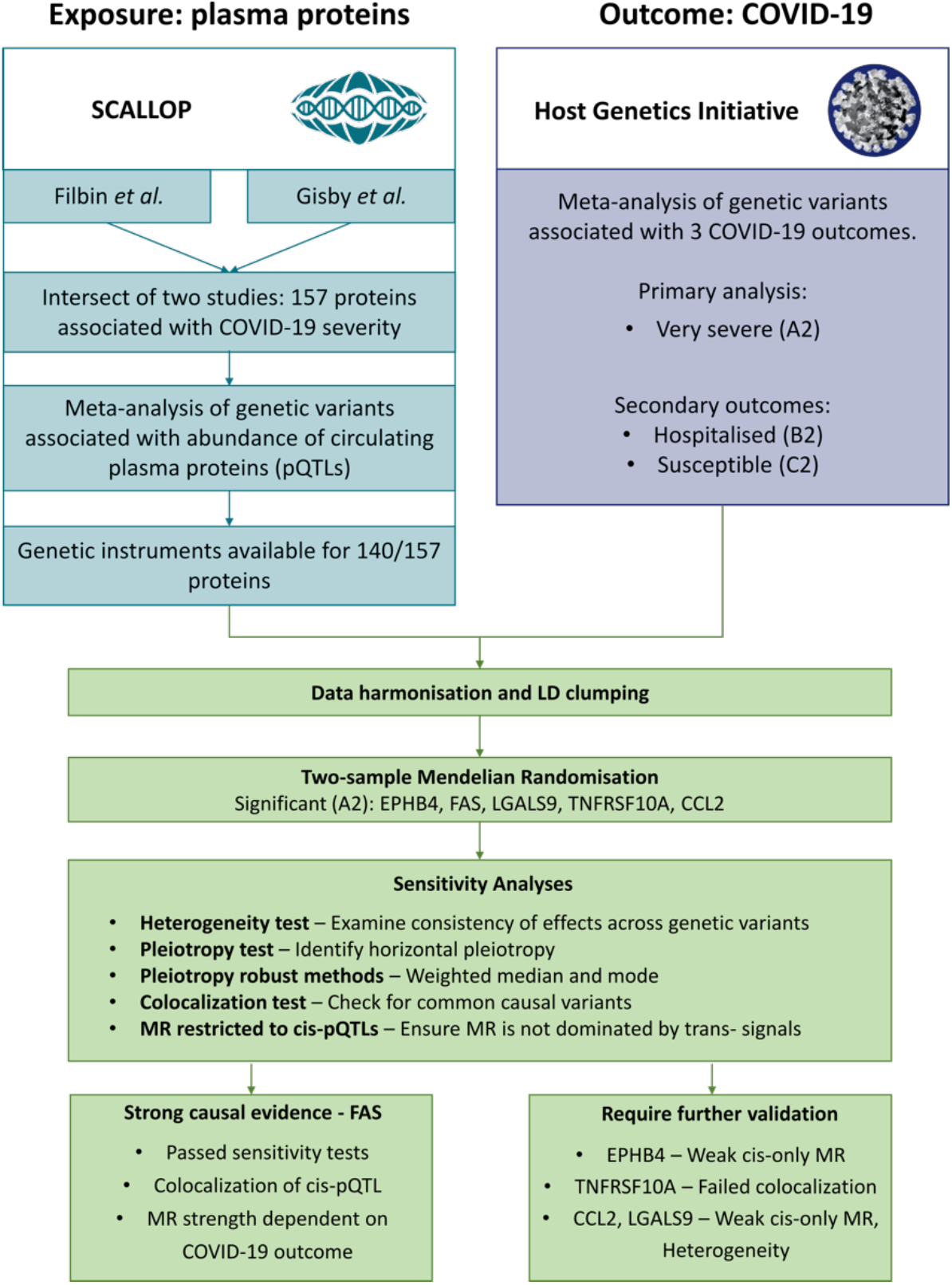
Mendelian randomisation study design and data sources. Severity-associated protein biomarkers were identified from the studies by Filbin *et al*^*10*^ and Gisby *et al*^*11*^.

To identify genetic instruments for MR, we accessed data from large European-heritage meta-analyses of plasma pQTL studies that also used the Olink platform performed by the SCALLOP consortium (https://www.olink.com/scallop/)^15^. The sample sizes of the protein GWAS meta-analyses varied from 3,658 to 26,494 (**Methods**). MR analysis was possible for 140 proteins, where at least one non-HLA pQTL was available. For 30 proteins there were only local-acting (‘cis’) pQTL (i.e., the pQTL lies within +/-0.5 Mb of the gene encoding the protein), for 17 there were only distant-acting (‘trans’) pQTL, and for 93 there were both. At each pQTL, we performed linkage disequilibrium pruning (LD r^2^≤0.01) to remove correlated variants, prior to MR (**Methods**).

For the outcome data, we used GWAS summary statistics from the COVID-19 Host Genetics Initiative release 5 (HGI: https://www.covid19hg.org)^17^ for very severe respiratory COVID-19 (defined as hospitalised patients requiring respiratory support and/or who died (analysis A2); for brevity, hereafter referred to as ‘severe COVID-19’).

We identified five proteins, EPH receptor B4 (EPHB4), C-C motif chemokine ligand 2 (CCL2), galectin 9 (LGALS9), Tumour Necrosis Factor Receptor Superfamily Member 10A (TNFRSF10A) and Fas cell surface death receptor (FAS), with significant MR causal estimates for severe COVID-19 after multiple testing correction (5% false discovery rate (FDR)) (**Table 1**).

**Table 1:** Mendelian Randomisation of COVID-19 severity-associated circulating proteins and risk of severe COVID-19.

| Protein* | # IV | MR P | FDR | OR (95% CI) | cis OR (95% CI) | cis P | Cochran's Q (p-value) | Egger Intercept P |
| --- | --- | --- | --- | --- | --- | --- | --- | --- |
| EPHB4 | 5 | $1.28 \times 10^{-6}$ | $1.69 \times 10^{-4}$ | 0.56<br>(0.44-0.71) | 0.64<br>(0.36-1.15) | 0.13 | 4.3<br>(0.36) | 0.90 |
| CCL2 | 7 | $2.43 \times 10^{-6}$ | $1.69 \times 10^{-4}$ | 1.54<br>(1.29-1.84) | 0.71<br>(0.26-1.92) | 0.50 | 97.0<br>($1.1 \times 10^{-18}$) | 0.98 |
| LGALS9 | 26 | $6.38 \times 10^{-4}$ | $2.96 \times 10^{-2}$ | 0.79<br>(0.69-0.91) | 0.90<br>(0.78-1.04) | 0.16 | 54.7<br>($5.4 \times 10^{-4}$) | 0.05 |
| TNFRSF10A | 28 | $1.71 \times 10^{-3}$ | $4.76 \times 10^{-2}$ | 0.81<br>(0.71-0.92) | 0.81<br>(0.71-0.92) | $1.91 \times 10^{-3}$ | 9.4<br>(0.36) | 0.03 |
| FAS | 7 | $1.36 \times 10^{-3}$ | $4.74 \times 10^{-2}$ | 1.40<br>(1.14-1.72) | 1.44<br>(1.17-1.78) | $6.70 \times 10^{-4}$ | 29.1<br>(0.15) | 0.05 |

An important assumption of MR is that genetic instruments (here pQTL) affect the outcome (here severe COVID-19) only through the exposure (here the protein), and not through observed or non-observed confounding factors (the ‘no horizontal pleiotropy’ assumption)^18,19^. We therefore used a multi-layered strategy to assess whether our results were robust (**Methods, Fig. 1**). First, we used heterogeneity tests to test whether there was consistency in the causal effect estimates across the genetic variants used. Second, we performed sensitivity analyses using alternative MR methods that are robust to horizontal pleiotropy, including MR Egger, weighted mode and median methods. Third, we performed MR restricting genetic instruments to cis-pQTL. Finally, we used colocalization to test whether the pQTL and severe COVID-19 genetic association signals reflected the same or distinct underlying causal variants.

For 3 proteins (EPHB4, FAS and TNFRSF10A) there was no heterogeneity in effect estimates between individual genetic variants, and effect estimates of pleiotropy-robust methods were similar to those of the inverse-variance method. In contrast, for LGALS9 and CCL2, we observed significant heterogeneity (Cochran’s Q >50 and heterogeneity p-value <0.05) (**Table 1, Supplementary Fig. 1a)**, casting doubt on the reliability of the causal inference for these two proteins.

Cis protein quantitative trait loci (cis-pQTL), genetic variants that lie near the gene encoding the affected protein, are considered to be more reliable MR instruments since their direct relationship with the protein means they are less likely to violate the ‘no horizontal pleiotropy’ assumption than trans-pQTL, which may act through indirect pathways^20^. Therefore, as an additional sensitivity analysis, we tested whether we observed consistent causal effects when limiting genetic instruments to cis-pQTL. This cis-only MR analysis revealed significant results for FAS (P 6.7×10^− 4^) and TNFRSF10A (P 1.9×10^−3^), but not EPHB4, CCL2 or LGALS9 (P>0.1); **Table 1, Supplementary Fig. 1b**).

Next, to disentangle causal relationships from confounding by linkage disequilibrium, we performed colocalisation analysis^21^ at each locus to test whether the same causal variant underlies the pQTL and the association with severe COVID-19. For the cis-pQTL for FAS, there was convincing colocalisation of the pQTL and the severe COVID-19 signal (posterior probability (PP) of shared causal variant 0.95) (**Fig. 2a**). In contrast, for the cis-pQTL for TNFRSF10A, it was clear that the pQTL and the disease association were driven by different causal variants (PP of distinct causal variants 0.87, **Supplementary Fig. 2a**). Thus, we have evidence to support a causal role for FAS, but not TNFRSF10A, in severe COVID-19.

**Figure 2:**
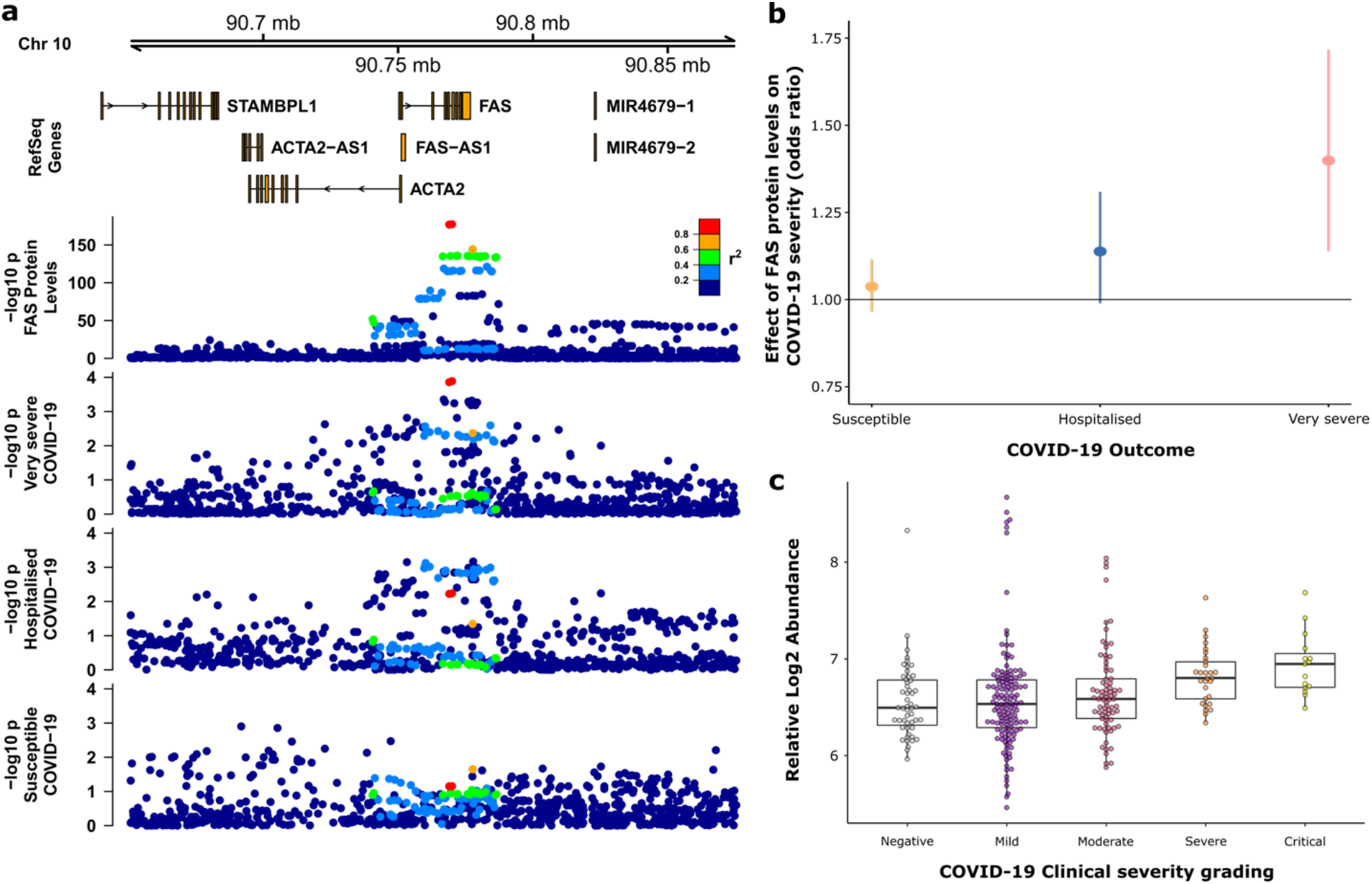
Mendelian Randomisation (MR) of soluble FAS protein levels and COVID-19 outcomes. **a)** Regional association plot (hg19 genome build) showing the cis-pQTL for soluble FAS (plasma) and the associations with COVID-19 outcomes. Posterior probability of a shared causal variant (PP H4) between FAS protein levels and very severe COVID-19 = 0.95. **b)** MR estimates of the causal effect of soluble FAS protein on different COVID-19 outcomes: susceptibility to infection, hospitalisation and very severe disease. **c)** Soluble FAS protein levels in COVID-19 patients, stratified by clinical severity, and non-infected controls (data from Gisby *et al*.^11^). Boxplots showing distribution of plasma protein levels according to COVID-19 status at the time of blood draw. Boxplots indicate median and interquartile range. n=256 samples from 55 COVID-19 patients and 51 samples from non-infected patients. ‘COVID-19 status’ indicates clinical severity score of the patient at the time the sample was taken. Mild n=135 samples; moderate n=77 samples; severe n=29 samples; critical n= 15 samples.

To empirically evaluate whether there was evidence of horizontal pleiotropy, we examined whether the cis genetic instruments for FAS used in the MR analysis, or variants in LD with them (r^2^>0.6), were associated with any protein. We utilised the PhenoScanner database that contains >5,000 genotype-phenotype associations^22,23^ and found no other associations with other proteins (at P <1×10^−5^).

To validate our results, we repeated the two-sample MR for soluble FAS in severe COVID-19 using genetic instruments derived from an independent pQTL dataset from a study using an alternative proteomics platform, the aptamer-based Somascan^*14*^. Consistent with our primary analysis, this revealed that genetic predisposition to higher circulating soluble FAS levels is associated with increased risk of severe COVID-19 (MR estimate OR = 1.35 [95% CI 1.14-1.60], p-value = 4.6×10^−4^) (**Supplementary Fig. 3**), providing independent support for our findings.

Having established evidence for a putatively causal role for FAS in severe COVID-19, we asked whether it may also play a role in susceptibility to COVID-19. We repeated the MR analysis using different COVID-19 GWAS datasets: all individuals with COVID-19 versus controls (i.e. susceptibility to COVID-19 – HGI C2 analysis), and all hospitalised COVID-19 patients vs controls (i.e. selecting for a degree of severity – HGI B2 analysis). Strikingly, we saw a gradient of MR effects across these outcomes. FAS showed no causal effect on susceptibility to COVID-19, a weak effect on COVID-19 hospitalisation and strong effect on severe COVID-19 (**Fig. 2b**). These data suggest that genetic propensity to higher soluble FAS levels influences COVID-19 severity, but not susceptibility. Observational data from the analysis of Gisby *et al*.^11^ revealed a similar pattern. Plasma FAS was not significantly differentially abundant in the comparison of all COVID-19 cases versus uninfected controls (Benjamini-Hochberg adjusted P value 0.280), but it was highly significantly associated with COVID-19 severity grading within-cases (Benjamini-Hochberg adjusted P 0.019) (**Fig. 2c**).

We next investigated the mechanism underlying the cis-pQTL for plasma FAS. The most strongly associated pQTL variant for FAS was rs982764 (**Supplementary Table 1**), an intronic single nucleotide polymorphism (SNP) in *FAS*, located between exons 4 and 5. This is a common variant, with minor allele frequency (MAF) ∼31% in European ancestry individuals. The major allele, rs982764:T, was associated with higher circulating soluble FAS levels and higher risk of severe COVID-19 (**Supplementary Fig. 4)**. The sentinel variant, rs982764, was not in LD (r2 >0.2) with any non-synonymous protein-coding variants. We therefore evaluated whether the pQTL was mediated through regulation of gene expression by examining eQTL data from the GTEx Catalogue (multiple tissues), whole-blood data from eQTLGen^24^, and sorted immune cell subsets from Peters *et al*.^*25*^. While these data revealed a cis-eQTL for FAS that was common to multiple tissues (e.g. lung, whole-blood, monocytes, adipose tissue, and artery), the eQTL signal did not colocalise with the pQTL (PP of distinct causal variants of 1.00 for both eQTLGen and GTEx) (**Supplementary Fig. 5**). Since the results of colocalisation methods can be affected by the presence of multiple independent variants, we then performed colocalisation using a method that allows for multiple causal variants (SuSiE^26^). This confirmed that, even after accounting for conditionally independent signals, the eQTL signal and pQTL signals for FAS did not colocalise (PP of distinct causal variants = 1.00).

Given that the pQTL could not be explained by an eQTL, we hypothesised that it might arise due to genetically influenced alternative splicing. FAS is a receptor for the cytokine FAS-ligand (FASL), and binding of FASL to FAS on the cell surface triggers an intracellular signalling cascade that leads to apoptosis of the cell. In humans, the *FAS* gene has 9 exons and encodes a cell surface receptor consisting of an extracellular portion, a transmembrane domain and an intracellular portion. In addition to the full-length *FAS* mRNA, several shorter transcripts arising from alternative splicing have been described (**Fig. 3a**)^27,28^. Alternative splicing leading to skipping of exon 6 (transcript isoform *FASΔEx6*) results in a secreted protein lacking the transmembrane domain^28^. This soluble form of FAS acts as a decoy receptor for FASL and thus has biologically opposing actions to membrane-bound FAS by reducing FAS-FASL signalling, resulting in reduced apoptosis^29^.

**Figure 3:**
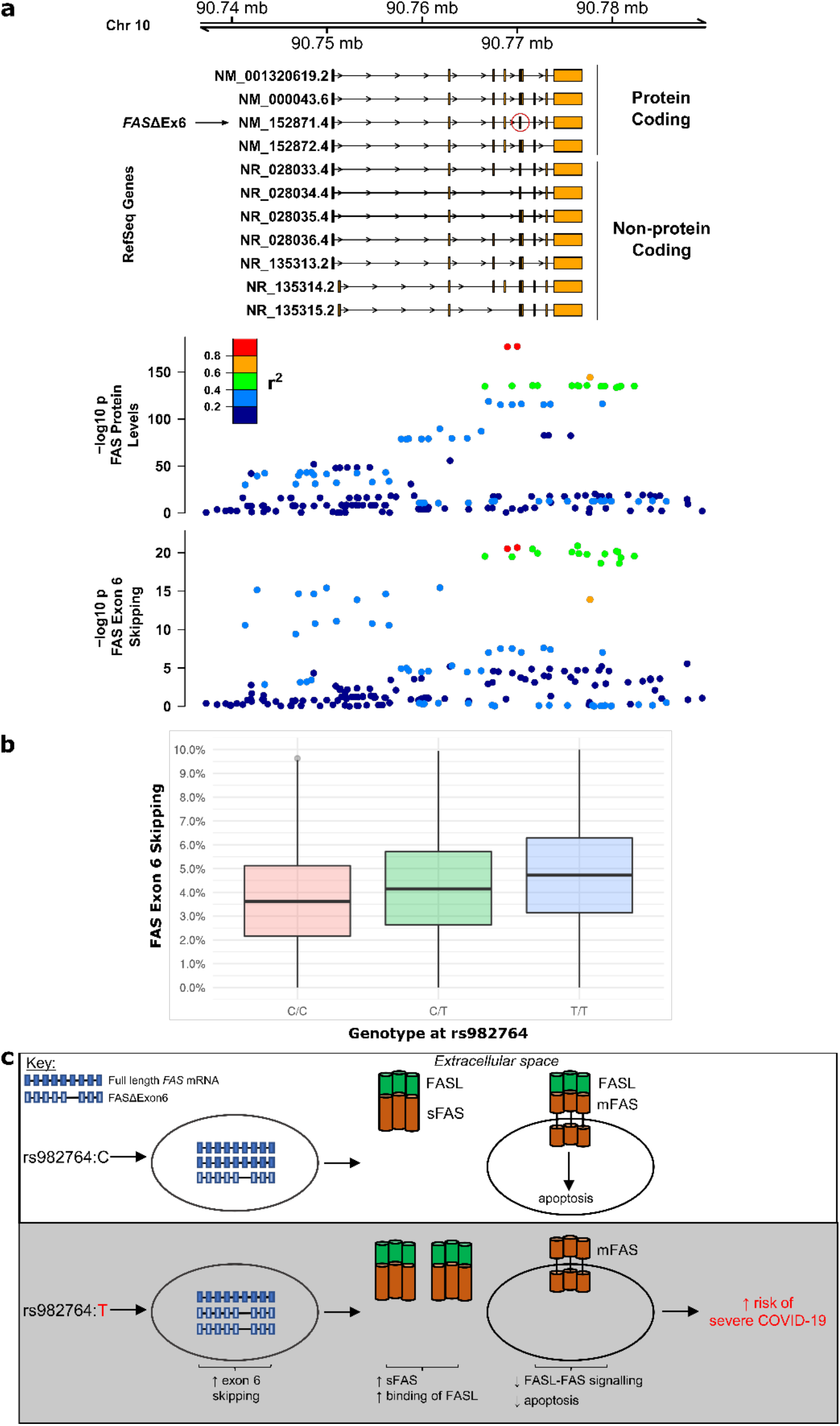
**a)** Regional association plot showing (from top to bottom): transcript isoforms, the soluble FAS cis-pQTL, and the associations with FAS exon 6 splicing. *FASΔEx6* indicates the transcript isoform lacking exon 6 (red circle). The posterior probability of a shared causal variant (PP H4) between FAS protein levels and exon 6 splicing = 0.99. **b)** Boxplot showing exon 6 splice quantitative trait locus (sQTL). Number of individuals by genotype at rs982764: 443 (CC), 1992 (CT), 2329 (TT). In 14 individuals genotype could not be reliably ascertained. Y-axis represents % of transcripts with exon 6 skipping. P for association with genotype 4×10^−22^ (linear model). **c)** Proposed model by which genetic variation in *FAS* increases risk of severe COVID-19. A non-coding variant acts as a splice QTL. The risk allele for very severe COVID-19 (rs982764:T) is associated with an increased proportion of transcripts lacking exon 6 resulting in higher levels of soluble FAS (sFAS). sFAS acts as a decoy receptor for FAS-ligand (FASL), blocking FASL binding to membrane-bound FAS (mFAS) on the cell surface and thus reducing apoptosis.

We therefore tested the hypothesis that the pQTL for plasma FAS resulted from genetically regulated skipping of exon 6. Using whole-blood RNA-seq data from 4,778 individuals, we examined alternative splicing and tested for associations between variants in the *FAS* region and transcripts lacking exon 6. The sentinel pQTL SNP, rs982764, was strongly associated with exon 6 skipping (P= 2.1 × 10^−22^). Moreover, the exon 6 splice QTL displayed the same association pattern as the pQTL and the GWAS signal for very severe COVID-19 (**Fig. 3a**). Formal colocalisation analysis confirmed that the splice QTL and the pQTL were highly likely to reflect the same underlying causal variant (PP of shared causal variant of 0.998). The risk allele for severe COVID-19 (rs982764:T) was associated with a shift towards transcripts lacking exon 6, which are known to encode soluble FAS (**Fig. 3b**). We confirmed this empirically via our pQTL data, with rs982764:T associated with higher plasma soluble FAS abundance. Together these data reveal a novel genetic mechanism by which a non-coding variant impacts the risk of severe COVID-19 through alternative splicing leading to elevated soluble FAS.

## Discussion

Here we performed Mendelian Randomisation (MR) to evaluate whether proteins observationally associated with severe COVID-19 play a causal role in severe disease. We focused on severe COVID-19 as this is responsible for the loss of life and has threatened to overcome the capacity of healthcare systems across the world. In addition to vaccination programmes, there is an urgent need for therapies to ameliorate severe disease. This requires improved understanding of the aberrant host immune response in this subset of patients.

We took a broad, but hypothesis-driven approach, by focussing on proteins that have been shown to robustly associate observationally with disease severity in COVID-19 patients in two independent clinical cohorts. A strength of our study was the use of a large-scale pQTL meta-analyses to provide robust genetic instruments. Our MR analysis revealed that the genetic tendency to higher plasma soluble FAS increased the risk of very severe COVID-19, implicating soluble FAS as a causal factor in severe COVID-19. Using a range of COVID-19 patient severity phenotypes (any diagnosis of COVID-19 infection, hospitalisation due to COVID-19, and very severe COVID-19 requiring respiratory support) in our MR analyses revealed a gradient of causal effect size estimates proportional to COVID-19 severity (**Fig. 2b**). These data suggest that the FAS pathway plays a role specifically in the pathogenesis of severe disease, rather than susceptibility to COVID-19 infection. Examination of soluble FAS levels in the plasma of patients with COVID-19 revealed a similar pattern (**Fig. 2c**).

The *FAS* gene encodes the FAS death receptor, also known as tumour necrosis factor receptor superfamily member 6 (TNFRSF6). Alternative splicing leads to multiple transcript isoforms (**Fig. 3a**)^27,28^. The canonical isoform encodes a type 1 transmembrane protein that is the cell surface receptor for the cytokine FAS-ligand (FASL), and plays important roles in the control of apoptosis, particularly in lymphocytes^30^. Binding of FASL to the extracellular portion of membrane-bound FAS triggers an intracellular cascade resulting in apoptosis of the FAS-expressing cell. Apoptosis is mediated by a ‘death domain’, an 85 amino acid-long structure in the intracellular portion of FAS, encoded by exon nine^31^. In contrast, an isoform arising from skipping of exon 6 encodes a secreted protein lacking the transmembrane domain. This soluble form of FAS acts as a decoy receptor for FASL and therefore has biologically opposing actions to membrane-bound FAS, by reducing FAS-FASL signalling and thus blocking the pro-apoptotic pathway^29^.

Investigation of the mechanism underpinning the cis-pQTL for FAS revealed that the risk allele for severe COVID-19 influences FAS mRNA splicing, resulting in a greater proportion of transcripts lacking exon 6 (**Fig. 3b**), which in turn lead to more soluble FAS (**Supplementary Fig. 4**), the anti-apoptotic decoy receptor for FASL. Our data therefore support a model whereby genetic predisposition to reduced FASL signalling through membrane-bound FAS leads to increased risk of severe COVID-19 (**Fig. 3c**). We hypothesise that this may result in impaired apoptosis of activated lymphocytes (enhancing immune-mediated pathology) or virus-infected cells (retarding viral clearance). *In vitro*, treatment with ibrutinib, a Bruton’s tyrosine kinase (BTK) inhibitor used in the treatment of haematological malignancy, has been shown to decrease soluble FAS, thereby enhancing FAS-mediated apoptosis^32^, suggesting a potential repurposing opportunity for severe COVID-19. Interestingly, case reports describe clinical improvement in haematology patients with severe COVID-19 on re-instigation of ibrutinib therapy^33,34^.

*Fas* knock-out mice develop an autoimmune disease similar to human lupus, with anti-nuclear antibodies, nephritis, lymphadenopathy and splenomegaly^35^. Mirroring this, deleterious mutations in the *FAS* gene in humans result in a rare Mendelian disease (autoimmune lymphoproliferative syndrome, ALPS, OMIM: 601859), characterised by autoimmunity and lymphoproliferative disease as a result of defective lymphocyte apoptosis^36-38,39^. In addition, common variants in the *FAS* gene region are associated with the proportion of lymphocytes in the blood white cell count^40^, chronic lymphocytic leukaemia (CLL)^41,42^, and autoimmune diseases including juvenile idiopathic arthritis (JIA)^43^. The risk allele for JIA (rs7069750:C) reduces FAS gene expression.

These observations reveal striking parallels in the spectrum of immune-mediated disease phenotypes related to genetic variation in *FAS*. Non-functional FAS protein (resulting from rare coding mutations) and quantitatively reduced levels of FAS gene expression (due to common non-coding polymorphisms) both predispose to autoimmune disease. Similarly, we show that elevation of soluble FAS, which inhibits signalling via membrane-bound FAS, increases susceptibility to severe COVID-19. Thus distinct genetic variants in the *FAS* gene converge on impaired FASL-FAS signalling, and result in immunopathology.

Other studies have performed Mendelian randomisation of proteins in COVID-19^44,45^. The MR analysis of Zhou *et al* identified OAS1 as a causal factor common to COVID-19 susceptibility, hospitalisation and very severe disease^44^. In contrast, we identified FAS as contributing specifically to severe COVID-19, but not susceptibility to infection. Gaziano *et al*.^*45*^ identified IFNAR2 and ACE2 as playing causal roles in COVID-19.

In summary, we demonstrate that that genetic tendency to higher levels of soluble FAS is a causal factor in severe COVID. Moreover, we reveal a novel genetic mechanism by which the risk allele for severe COVID-19 influences susceptibility through alternative splicing of FAS. This non-coding variant affects alternative splicing, resulting in increased levels of soluble FAS, a decoy receptor for FASL which blocks signalling of FASL via membrane-bound FAS on the cell surface. Our data provide insights into the pathogenesis of severe COVID-19 and suggest a potential therapeutic opportunity from restoration of FASL signalling.

## Methods

### Mendelian Randomisation

Mendelian randomisation uses genetic variants as instrumental variables in order to investigate the effects of a risk factor (exposure) on a disease (outcome), provided certain assumptions hold. The method reduces bias created from confounding, by treating the variants used as equivalent to treatment allocation in randomized control trials^13,46-48^. We used two-sample Mendelian Randomization (MR) to test the causal role of plasma proteins in severe COVID-19.

### Defining a list of proteins robustly associated with COVID-19 severity

We identified plasma proteins that were significantly associated (5% FDR) with COVID-19 severity in the two studies that used Olink proteomics platform (Filbin *et al*.^*10*^ and Gisby *et al*.^*11*^). To define a set of robust COVID-19 biomarkers, we took the intersect of the lists of significant associations from these two studies. This resulted in 157 proteins.

### Identification of genetic instruments through pQTL mapping

To provide a set of genetic instruments for MR, we performed a meta-analysis of pQTL studies through framework of the SCALLOP consortium. All contributing pQTL studies had been performed using plasma with proteins measured using Olink immunoassays (Olink Bioscience, Uppsala, Sweden).

### Cohort-level pQTL analysis

Details of the cohorts and cohort-specific ethical approval are included in the **Supplementary Table 2**. Plasma protein levels were measured using up to 5 Olink 92-plex immunoassays (“Inflammation”, “Cardiovascular2”, “Cardiovascular3”, “Cardiometabolic” and “Immune Response”). Despite the nomenclature, inflammation and immune related proteins were highly enriched on all panels. The sample size per protein across all available cohorts varied from 3,658 (for proteins on the Immune Response panel) to to 26,494 (from proteins on the cardiovascular2 and cardiovascular3 panels). All subjects were of European heritage. Protein levels were rank-based inverse-normal transformed prior to genetic association testing. Genome-wide association analyses were performed using an additive regression model of protein on genotype with adjustment for age, sex and cohort specific covariates. Population structure was accounted for using by including principal components as covariates or by accounting for relatedness using linear mixed models as appropriate to the specific cohort.

### pQTL meta-analysis

Prior to meta-analysis cohort-level summary statistics were quality controlled EasyQC software^49^, following the protocol as described in Winkler *et al*.^*49*^. Meta-analysis was performed using inverse-variance fixed effect method implemented in METAL^50^ (‘STDERR’ option), followed by correction for genomic control. Meta-analysis summary statistics were then further filtered for minor allele frequency (MAF) > 0.01, and heterogeneity in effect size estimates. Variants with heterogeneity I^2^≥75% were not considered significant and were removed prior to further analysis.

### pQTL locus definition

To define the boundaries of each pQTL locus, we first selected all genetic variants with p-value<1×10^−5^ and then calculated the distance between each consecutive variant located on the same chromosome. Two consecutive variants were identified as belonging to different loci if they were more than 250 kb apart. The sentinel variant was defined as the variant with the lowest p-value within the locus. A locus was defined as a cis-pQTL if the sentinel variant was within 0.5 Mb of the start or end of the gene encoding the given protein, otherwise it was classified as a trans-pQTL.

### Outcome data for MR testing: COVID-19 GWAS

We accessed COVID-19 GWAS data from the COVID-19 Host Genetics Initiative website (https://www.covid19hg.org/). To match the ancestry of the individuals in the pQTL meta-analysis, we downloaded data from European-ancestry individuals. Specifically, we downloaded the following datasets: European summary statistics without 23 and me data from the HGI website release 5 (release date 7^th^ January 2021) for A2 (very severe respiratory confirmed COVID-19 versus population), B2 (hospitalised COVID-19 versus population), C2 (susceptibility - COVID-19 versus population). Prior to downstream analyses, all variants with heterogeneity p-value 0.001 (as per HGI recommendation) were removed. The following links were used to download the data: https://storage.googleapis.com/covid19-hg-public/20201215/results/20210107/COVID19_HGI_A2_ALL_eur_leave_ukbb_23andme_20210107.b37.txt.gz https://storage.googleapis.com/covid19-hg-public/20201215/results/20210107/COVID19_HGI_B2_ALL_eur_leave_ukbb_23andme_20210107.b37.txt.gz https://storage.googleapis.com/covid19-hg-public/20201215/results/20210107/COVID19_HGI_C2_ALL_eur_leave_ukbb_23andme_20210107.b37.txt.gz

The goal of our MR analysis was to test the causal role of proteins in very severe COVID-19 and so for our principal analyses we used the A2 COVID-19 GWAS dataset as the outcome. For FAS, the significant protein identified by our principal MR analysis (FAS), we also performed MR using COVID-19 dataset B2 and C2.

### Details of MR testing

#### Primary MR analysis

For each protein, MR evaluating its causal role in very severe COVID-19 was performed using the TwoSampleMR package^51^. Where a single variant was used as the genetic instrument, we performed a Wald Ratio (WR) test. In the case of multiple genetic variants, we used the fixed effects inverse variance weighted (IVW) method.

#### Variant selection

For each protein first we selected genetic variants associated with the protein level at genome-wide significance (P-value < 5×10^−8^). From these, we retained variants that were also present in the outcome (very severe COVID-19) GWAS summary statistics. Next, to obtain a set of genetic instruments with low correlation, we performed LD pruning of these variants using Plink 1.9^52^ and the options clump_r2 = 0.01 and clump_kb = 10,000. The LD reference panel for the pruning procedure was created by randomly selecting 10,000 unrelated individuals of British ancestry (evaluated on the basis of genomic data) from UK Biobank, followed by removing positional and marker name duplicated SNPs from imputed genotypes using --rm-dup exclude-all function using Plink 2.0^52^. Variants in the MHC region (hg19 genome build chr6:26,000,000-34,000,000) were excluded.

#### Sensitivity analyses

Since MR relies on certain assumptions, that often cannot be directly evaluated, we performed a range of sensitivity analyses. We used statistical methods to test for horizontal pleiotropy including MR Egger^53^. We performed MR using the weighted mode and median methods^54^, which are more robust to the presence of horizontal pleiotropy, and the maximum likelihood (ML) method. The ML method allows for uncertainty in the effect size of the genetic associations with the exposure (unlike the IVW method which uses simple weights), and it allows for genetic associations with the exposure and with the outcome for each variant to be correlated^55^.

#### Cis-only MR

As additional sensitivity analysis we repeated MR restricting the genetic instruments to cis-pQTLs only, as these are less likely to be affected by horizontal pleiotropy^20^. To select cis-only instruments we performed LD pruning (as described earlier) on all variants with p<5×10^−8^ within a cis locus.

## Replication of FAS MR

The genetic instruments used in the primary analysis were pQTL identified in a meta-analysis of studies that used Olink immunoassays. To validate this, we repeated the MR analysis for FAS using an alternative pQTL dataset based on proteomic profiling using the aptamer-based SomaLogic platform^14^. The COVID-19 GWAS data was the same as for the primary analysis (HGI A2 GWAS summary statistics).

### Colocalisation

To distinguish causal relationships from confounding by LD we used colocalisation analysis. Colocalisation analysis tests whether regional genetic association signals for different traits arise from distinct or the same shared causal variant. The Bayesian colocalisation method implemented in the coloc.abf() function from the “coloc”^21^ R package provides posterior probabilities (PP) for 5 different hypotheses: the null hypothesis of no association with either of the traits (H0) and four alternative hypotheses of either association with only one of the traits (H1, H2), or association of both traits but under the effect of distinct underlying causal variants (H3), or association of both traits under the effect of a shared causal variant (H4) i.e. colocalisation. For candidate proteins identified by the MR analysis, we compared the HGI A2 COVID-19 regional association signal(s) with that of the relevant pQTL. We considered a PP>0.8 as a strong evidence in favour of that hypothesis.

We also used coloc to test whether the FAS cis-pQTL colocalised with eQTLs from multiple cell types across multiple studies. These included multiple tissue types from GTEx v7 (obtained from the GTEx Portal on 03/23/21), whole blood data from eQTLGen^24^, and 5 sorted leukocyte subsets from Peters *et al*.^*25*^. An assumption of coloc is that there is at most one causal variant at the locus per trait. To allow for the possibility of multiple independent eQTLs or pQTLs, we used the Sum of Single Effects method^26^, which allows for simultaneous colocalisation testing of multiple causal variants.

### FAS levels in COVID-19 patients

We used data on COVID-19 patients and sex, age and ethnicity matched non-infected controls from the study by Gisby *et al*^*11*^. The study design involved serial plasma sampling from the COVID-19 patients. For full details of association testing see Gisby *et al*^*11*^. Briefly, differential abundance analysis of FAS levels between COVID-19 patients and controls was performed using linear mixed models with age, sex and ethnicity as covariates. Associations of FAS and clinical severity scores were performed within COVID-19 cases, using a 4-level ordinal scale for clinical severity (mild, moderate, severe and critical) at the time of blood sampling. Again, linear mixed models were used to account for repeated measurements from the same individual.

### RNA-sequencing and splice QTL analysis

In 5,000 individuals of the INTERVAL cohort, 3 ml of whole blood were collected in Tempus Blood RNA Tubes (ThermoFisher Scientific), following the manufacturer’s instructions. RNA extraction was performed by QIAGEN Genomic Services using an in-house developed protocol based on QIAGEN’s proprietary silica technology. We assessed the quality of the extracted RNA using spectrophotometric analysis. RNA Integrity Number (RIN) values were determined using a TapeStation 4200 system (Agilent), following the manufacturer’s protocol. Messenger RNA (mRNA) was isolated using a NEBNext Poly(A) mRNA Magnetic Isolation Module (NEB). Globin depletion was performed using a KAPA RiboErase Globin Kit (Roche). RNA library preparation was done using a NEBNext Ultra II DNA Library Prep Kit for Illumina (NEB) on a Bravo WS automation system (Agilent). Libraries were pooled to 96-plex in equimolar amounts, quantified using a High Sensitivity DNA Kit on a 2100 Bioanalyzer (Agilent), and then normalised to 2.8 nM prior to sequencing. Samples were sequenced using 75 bp paired-end sequencing reads (reverse stranded) on a NovaSeq 6000 system (S4 flow cell, Xp workflow; Illumina). We assessed the sequence data quality using FastQC v0.11.8. Reads were aligned to the GRCh38 human reference genome (Ensembl GTF annotation v99) using STAR v2.7.3.a^56^. Data from 4,778 individuals were subjected to downstream analyses. We extracted transcript splice junctions with the regtools “junction extract” tool, and introns were clustered and excision ratios calculated and normalised for downstream sQTL analysis according to the leafcutter^57^ pipeline (build #aa12b1e) with default parameters. Cis-sQTLs were calculated on the resulting leafcutter ratios for the excision event of FAS exon 6 (ENSE00003500194) and flanking introns, and genotypes with a minor allele frequency >0.01 in the region +/- 200kb of the FAS gene using tensorQTL^58^ 1.0.5. Blood cell type proportions, sex, and 10 genomic and 10 splicing principal components were added as covariates to the linear model.

## Data Availability

All data described in the manuscript is included in the Tables and Supplementary files.

## Supplementary Material

### Supplementary Tables and Figures

**Supplementary Figure 1:**
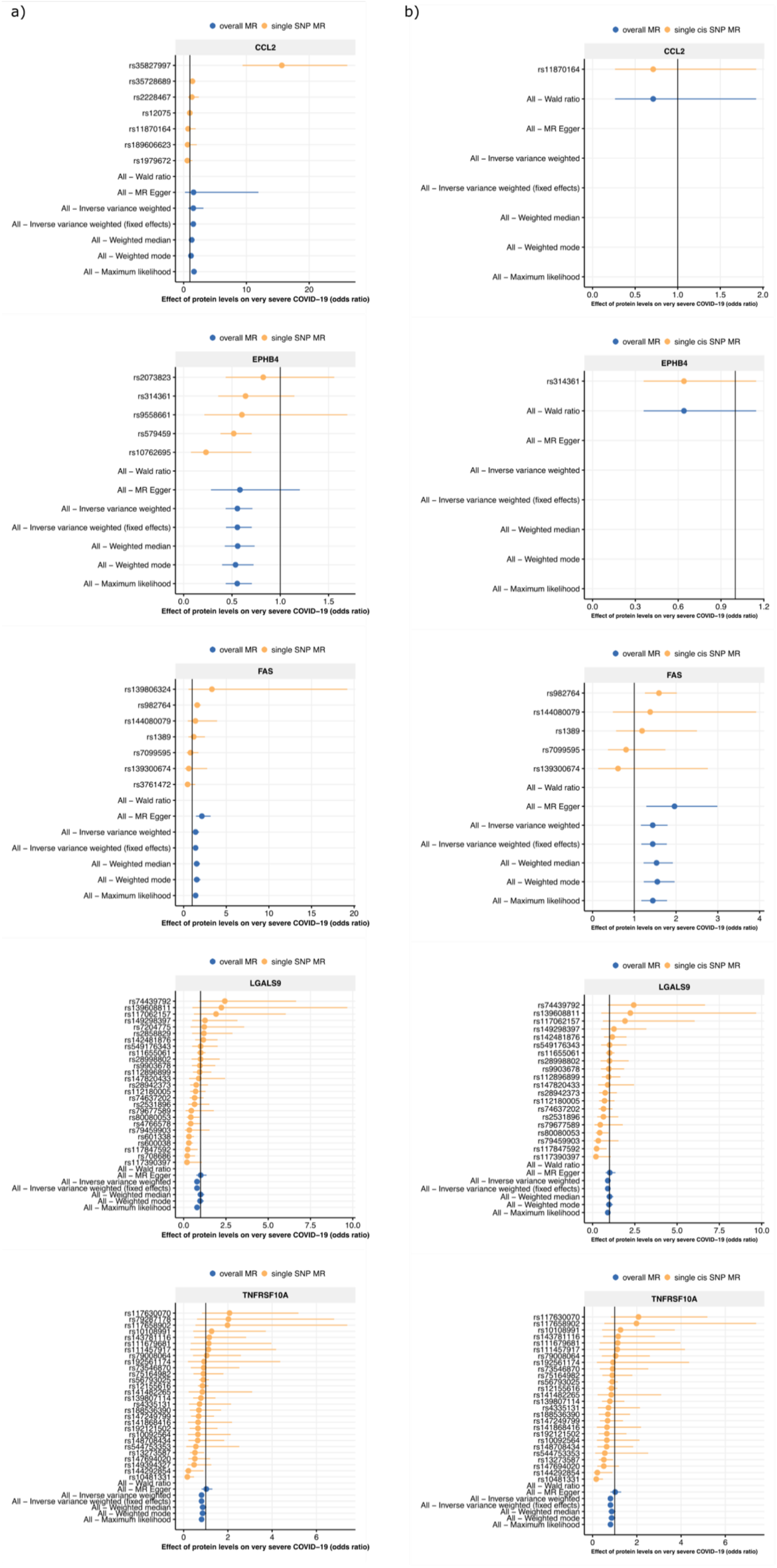
MR estimates using different MR methods for 5 proteins significant in the primary analysis (fixed effect inverse variance weighted method). a) MR effects estimated from all variants (both cis and trans). b) MR effects estimated using only cis variants. Vertical line represents odds ratio of 1.

**Supplementary Figure 2:**
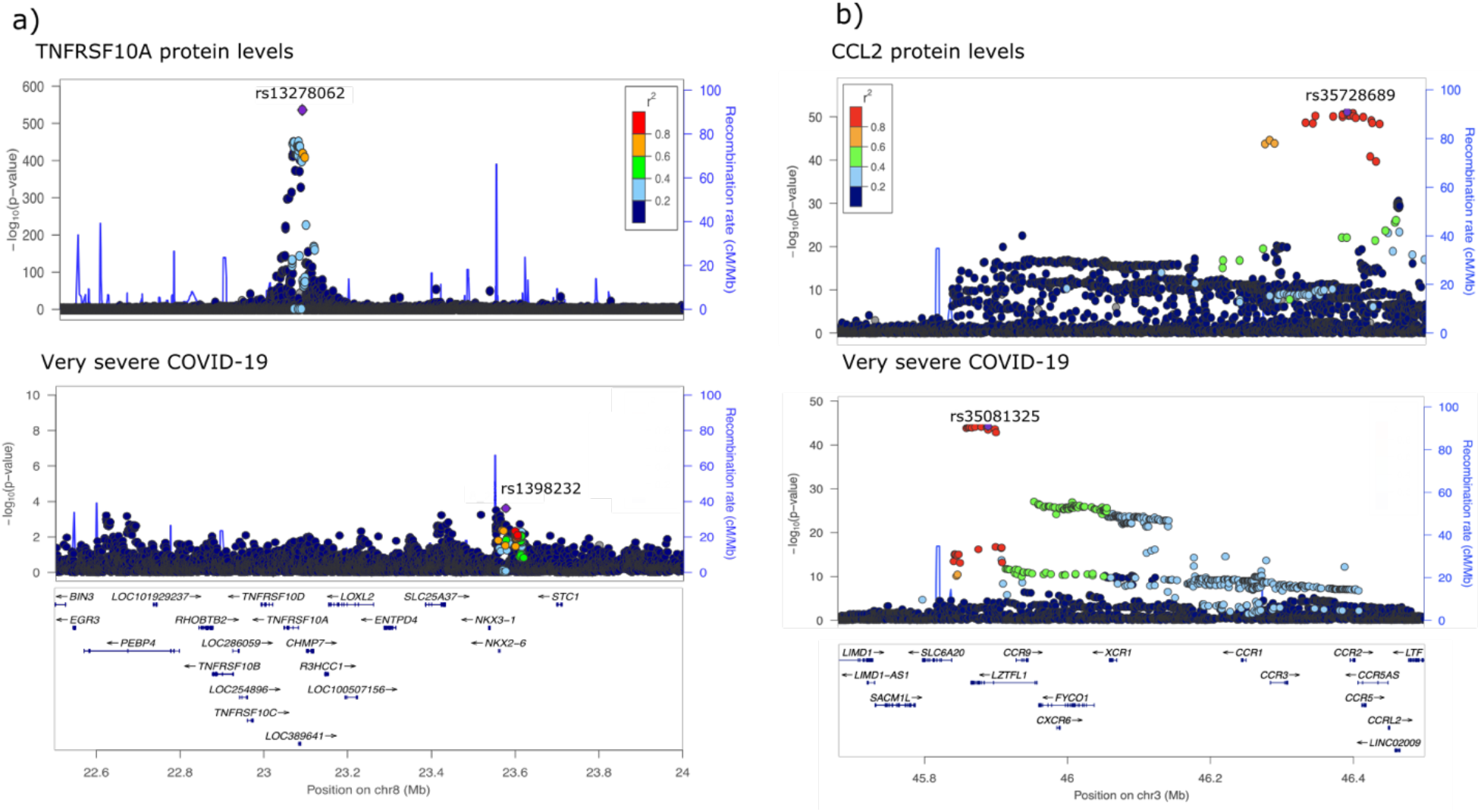
Genetic associations for TNFSRSF10A and CCL2 plasma protein levels and very severe COVID-19 do not colocalise. a) The sentinel pQTL (rs13278062) for TNFRSF10A is in linkage equilibrium with the sentinel COVID-19 variant (rs1398232). The posterior probability of distinct causal variants from colocalisation testing (PP H3) = 0.87. **b)** A trans-pQTL located on chromosome 3 (sentinel variant rs35728689) for CCL2 (which is encoded on chromosome 17) lies approximately 5kb upstream of the *CCR2* gene, which encodes the receptor for CCL2, suggesting an obvious biological mechanism for the trans-pQTL. Single-variant MR performed using this trans-pQTL was significant (rs35728689 P=2.8 ×10-2, Supplementary Table 1). However, LD r^2^ between sentinel CCL2 pQTL (rs35728689) and sentinel COVID-19 variant (rs35081325) is 0.006 in 1000 genomes EUR population and the posterior probability of distinct causal variants from colocalisation testing (PP H3) = 1.0.

**Supplementary Figure 3:**
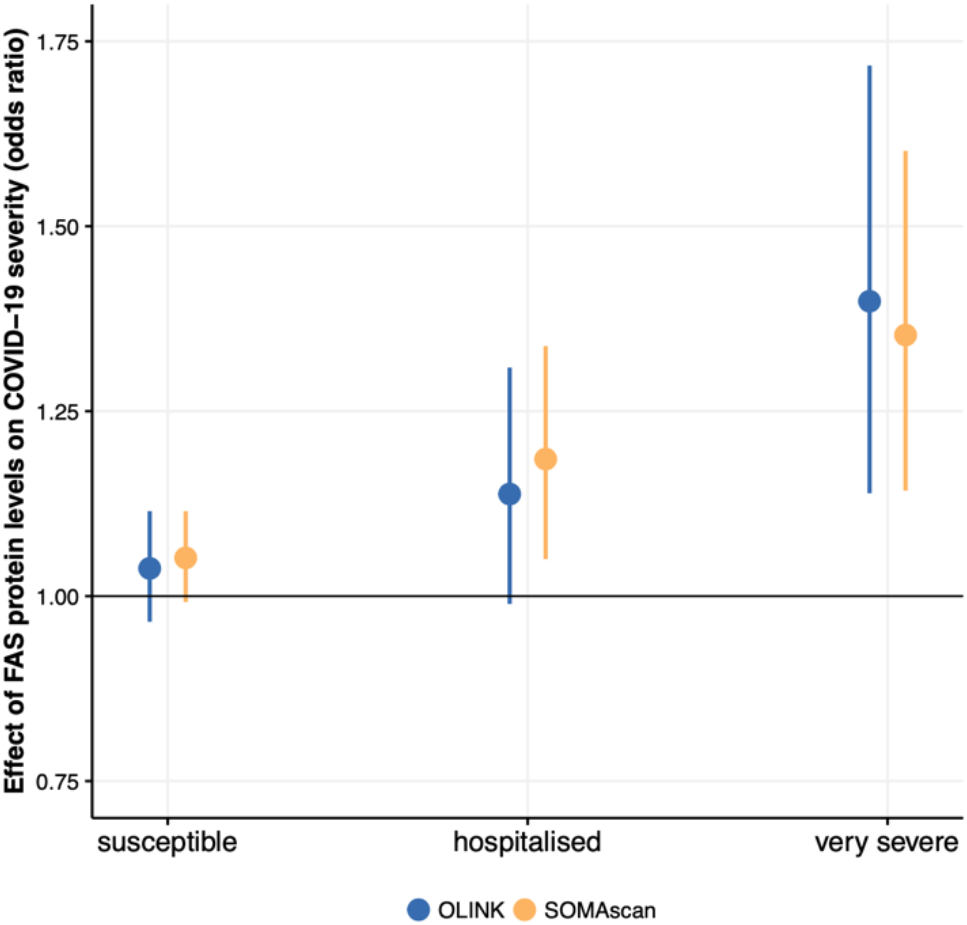
Validation of MR estimates of the causal effect of soluble FAS protein on COVID-19 outcomes. Genetic instruments were obtained from another pQTL study of FAS levels measured with a different proteomic platform (SOMAscan^14^). Details of individual instruments can be found in Supplementary Table 1. ‘Olink’ indicates the MR estimate obtained in our analysis.

**Supplementary Figure 4.**
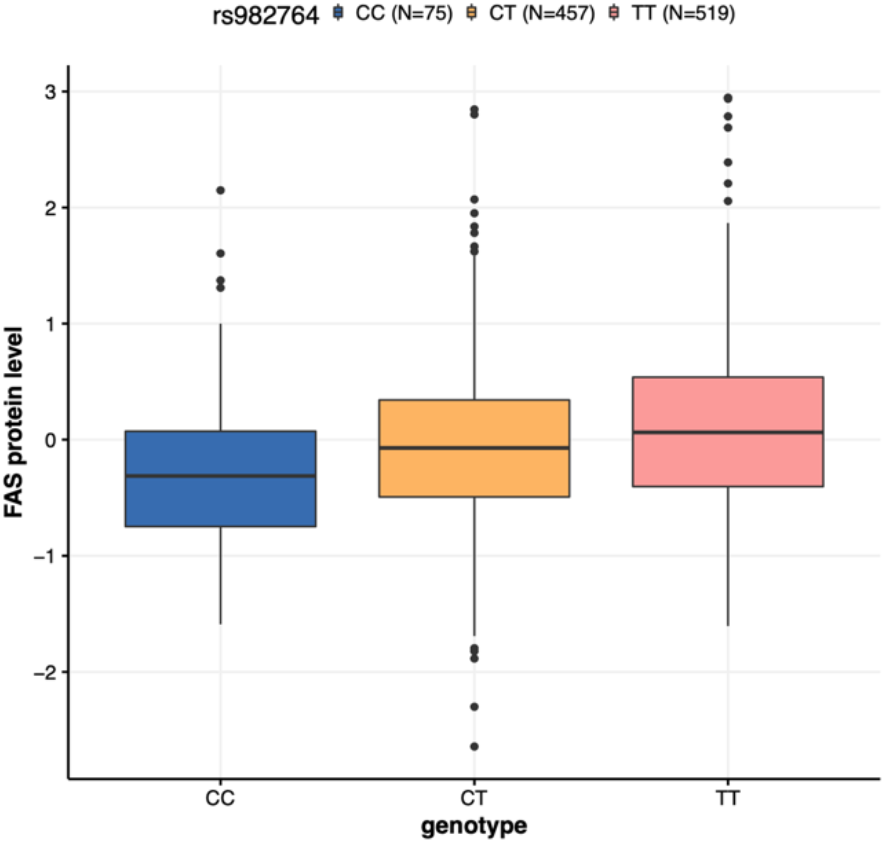
Boxplot showing genotype at rs982764 versus plasma soluble FAS levels (after correction for age, sex, batch effects, season of blood sampling and genetic principal components) in the ORCADES cohort.

**Supplementary Figure 5.**
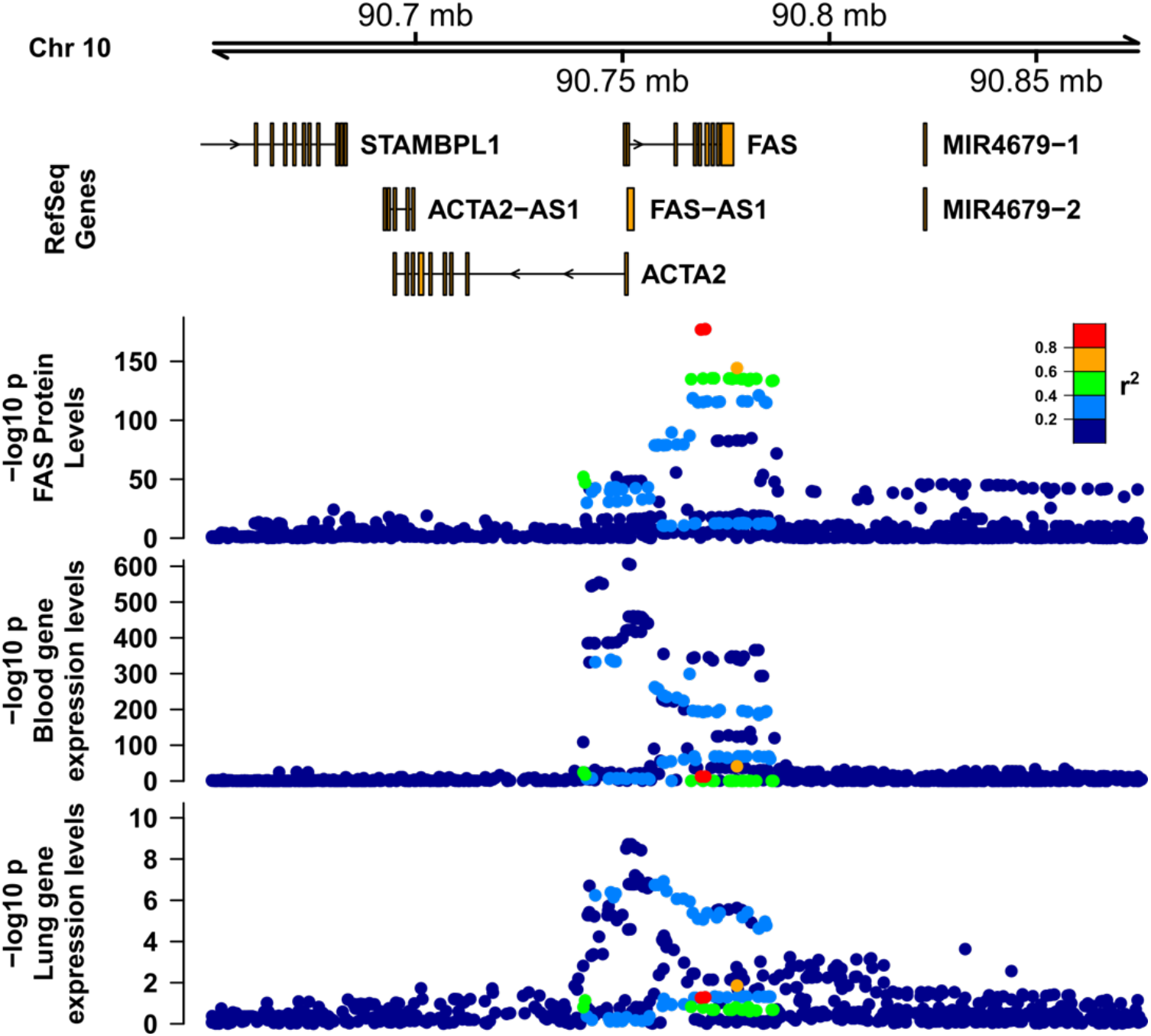
Regional association plots for FAS pQTL and eQTL. Tracks from top to bottom: FAS plasma pQTL (SCALLOP meta-analysis), FAS eQTL in whole blood (eQTLGen) and lung (GTex v7).

## Supplementary Tables (xlsx)

**Supplementary Table 1: SNP-wise estimates of the MR effect**. All - overall MR estimate, using all listed SNPs, exposure - protein levels, outcome - very severe COVID-19 (A2). genomic positions based on human genome build GRCh37

**Supplementary Table 2:** Details of contributing cohorts

## Acknowledgements

The proteomic work was carried out under the aegis of the SCALLOP consortium. We thank Aikaterini Siopi for administrative assistance. We thank the UK Biobank resource, approved under application 19655. Cohort-specific acknowledgements can be found in Supplementary Table 2. J.F.W. and C.H, acknowledge support from the MRC Human Genetics Unit programme grant “Quantitative traits in health and disease” (U.MC_UU_00007/10). J.E.P. acknowledges a UKRI Innovation Fellowship at Health Data Research UK (MR/S004068/2), a UKRI-DHSC COVID-19 Rapid Response Rolling Call (MR/V027638/1) and Community Jameel and the Imperial President’s Excellence Fund. The work of L.K. was supported by an RCUK Innovation Fellowship from the National Productivity Investment Fund (MR/R026408/1). J.H.Z. is funded by the NIHR Cambridge Biomedical Research Centre (BRC-1215-20014). A.L. is funded by the European Union’s Horizon 2020 research and innovation program IMforFUTURE, under H2020-MSCA-ITN grant agreement number 721815. B.P. is funded by a BHF Programme Grant (RG/18/13/33946). D.Z. was supported by the AHA Postdoctoral Fellowship [19POST34370115]. C.K. was supperted by NHLBI contracts 75N92021D00001, 75N92021D00002, 75N92021D00003, 75N92021D00004, 75N92021D00005. J.F. was supported by the Dutch Heart Foundation IN-CONTROL (CVON2018-27), the ERC Consolidator grant (101001678) and the Netherlands Organ-on-Chip Initiative, an NWO Gravitation project (024.003.001) funded by the Ministry of Education, Culture and Science of the government of The Netherlands’. N.M.C is a Wallenberg Center for Molecular Medicine Fellow. A.R. is funded by R01 HL136574 and S10OD02868. C.He. acknowledges the German Diabetes Center, (DDZ) which is funded by the German Federal Ministry of Health (Berlin, Germany), the Ministry of Culture and Science of the state North Rhine-Westphalia (Düsseldorf, Germany), and grants from the German Federal Ministry of Education and Research (Berlin, Germany) to the German Center for Diabetes Research e.V. (DZD). D.V.Z was supported by a VENI grant from NWO (194.006). J.G.S. was supported by grants from the Swedish Heart-Lung Foundation (2016-0134, 2016-0315 and 2019-0526), the Swedish Research Council (2017-02554), the European Research Council (ERC-STG-2015-679242), the Crafoord Foundation, Skåne University Hospital, the Scania county, governmental funding of clinical research within the Swedish National Health Service, a generous donation from the Knut and Alice Wallenberg foundation to the Wallenberg Center for Molecular Medicine in Lund, and funding from the Swedish Research Council (Linnaeus grant Dnr 349-2006-237, Strategic Research Area Exodiab Dnr 2009-1039) and Swedish Foundation for Strategic Research (Dnr IRC15-0067) to the Lund University Diabetes Center. J.D. holds a British Heart Foundation Professorship and a NIHR Senior Investigator Award. K.M. is supported by grants from the Swedish Research Council (grant no 2002-6147, 2005-6662, 2005-8214, 2008-2202, 2011-02427, 2015-03257, 2017-00644, 2017-06100 and 2019-01291, grants from the Swedish Research Council for Health, Working Life and Welfare (https://forte.se/en/; grant numbers 2011-0346, and 2017-00721), and annual grants from Avtal om Läkarutbildning och Forskning (ALF; Agreement concerning Cooperation on Medical Education and Research) between Uppsala University and Uppsala County Council. He acknowledges the national research infrastructure SIMPLER for provisioning of facilities and experimental support. SIMPLER receives funding through the Swedish Research Council under the grant no 2017-00644. A.T. was supported by the Wellcome Trust. K.S. was supported by the Biomedical Research Program at Weill Cornell Medicine in Qatar, a program funded by the Qatar Foundation, and by multiple grants from the Qatar National Research Fund (QNRF). C.L., E.W., and N.J.W. are funded by the Medical Research Council (MC_UU_12015/1). NJW is a NIHR Senior Investigator. R.J.S. is supported by a UKRI Innovation-HDR-UK Fellowship (MR/S003061/1). T.L.A was supported by NIDDK/NIH grant 1R01DK114183. J.K.B. acknowledges funding support from a BBSRC Institute Strategic Programme Grant (BBS/E/D/20002172), UKRI (MC_PC_20004, MC_PC_19025, MC_PC_1905, MRNO2995X/1) and the UK Intensive Care Society. RNA-sequencing in the INTERVAL study was supported by the Wellcome Trust (grant reference 206194/Z/17/Z), NHSBT and AstraZeneca. We thank the Sequencing Operations Team and Human Genetics Informatics Team (Guillaume Noell and Vivek Iyer) at the Wellcome Sanger Institute for performing the RNA library preparation and RNA-sequencing and analysis. EP is supported by the EU/EFPIA Innovative Medicines Initiative Joint Undertaking BigData@Heart grant 116074. We thank Dr Arianne Richard for helping with results interpretation and commenting on the manuscript.

## Disclaimer

The views expressed in this manuscript are those of the authors and do not necessarily represent the views of the National Heart, Lung, and Blood Institute; the National Institutes of Health; or the U.S. Department of Health and Human Services. The views expressed are those of the author(s) and not necessarily those of the NIHR or the Department of Health and Social Care.

## References

1. Szabo, P.A., et al. Longitudinal profiling of respiratory and systemic immune responses reveals myeloid cell-driven lung inflammation in severe COVID-19. Immunity (2021).

2. Wichmann, D., et al. Autopsy Findings and Venous Thromboembolism in Patients With COVID-19: A Prospective Cohort Study. Ann Intern Med 173, 268–277 (2020).

3. Carvalho, T., Krammer, F. & Iwasaki, A. The first 12 months of COVID-19: a timeline of immunological insights. Nature Reviews Immunology (2021).

4. Arunachalam, P.S., et al. Systems biological assessment of immunity to mild versus severe COVID-19 infection in humans. Science 369, 1210 (2020).

5. Interleukin-6 Receptor Antagonists in Critically Ill Patients with Covid-19. New England Journal of Medicine (2021).

6. Horby, P.W., et al. Tocilizumab in patients admitted to hospital with COVID-19 (RECOVERY): preliminary results of a randomised, controlled, open-label, platform trial. medRxiv, 2021.2002.2011.21249258 (2021).

7. Dexamethasone in Hospitalized Patients with Covid-19. New England Journal of Medicine 384, 693–704 (2020).

8. McInnes, I.B., Buckley, C.D. & Isaacs, J.D. Cytokines in rheumatoid arthritis — shaping the immunological landscape. Nature Reviews Rheumatology 12, 63–68 (2016).

9. Attwood, M.M., Jonsson, J., Rask-Andersen, M. & Schiöth, H.B. Soluble ligands as drug targets. Nature Reviews Drug Discovery 19, 695–710 (2020).

10. Filbin, M.R., et al. Plasma proteomics reveals tissue-specific cell death and mediators of cell-cell interactions in severe COVID-19 patients. bioRxiv : the preprint server for biology, 2020.2011.2002.365536 (2020).

11. Gisby, J., et al. Longitudinal proteomic profiling of dialysis patients with COVID-19 reveals markers of severity and predictors of death. eLife 10, e64827 (2021).

12. Hingorani, A. & Humphries, S. Nature’s randomised trials. The Lancet 366, 1906–1908 (2005).

13. Burgess, S., et al. Using published data in Mendelian randomization: a blueprint for efficient identification of causal risk factors. European journal of epidemiology 30, 543–552 (2015).

14. Sun, B.B., et al. Genomic atlas of the human plasma proteome. Nature 558, 73–79 (2018).

15. Folkersen, L., et al. Genomic and drug target evaluation of 90 cardiovascular proteins in 30,931 individuals. Nature Metabolism 2, 1135–1148 (2020).

16. Folkersen, L., et al. Mapping of 79 loci for 83 plasma protein biomarkers in cardiovascular disease. PLoS Genetics 13, 1–21 (2017).

17. The, C.-H.G.I. The COVID-19 Host Genetics Initiative, a global initiative to elucidate the role of host genetic factors in susceptibility and severity of the SARS-CoV-2 virus pandemic. European Journal of Human Genetics 28, 715–718 (2020).

18. Greenland, S. An introduction to instrumental variables for epidemiologists. International Journal of Epidemiology 29, 722–729 (2000).

19. Davey Smith, G. & Ebrahim, S. ‘Mendelian randomization’: can genetic epidemiology contribute to understanding environmental determinants of disease?*. International Journal of Epidemiology 32, 1–22 (2003).

20. Davey Smith, G. & Hemani, G. Mendelian randomization: genetic anchors for causal inference in epidemiological studies. Human Molecular Genetics 23, R89–R98 (2014).

21. Giambartolomei, C., et al. Bayesian Test for Colocalisation between Pairs of Genetic Association Studies Using Summary Statistics. PLOS Genetics 10, e1004383 (2014).

22. Staley, J.R., et al. PhenoScanner: a database of human genotype–phenotype associations. Bioinformatics 32, 3207–3209 (2016).

23. Kamat, M.A., et al. PhenoScanner V2: an expanded tool for searching human genotype–phenotype associations. Bioinformatics 35, 4851–4853 (2019).

24. Võsa, U., et al. Unraveling the polygenic architecture of complex traits using blood eQTL metaanalysis. bioRxiv, 447367 (2018).

25. Peters, J.E., et al. Insight into Genotype-Phenotype Associations through eQTL Mapping in Multiple Cell Types in Health and Immune-Mediated Disease. Plos Genetics 12(2016).

26. Wallace, C. A more accurate method for colocalisation analysis allowing for multiple causal variants. bioRxiv, 2021.2002.2023.432421 (2021).

27. Baeza-Centurion, P., Minana, B., Schmiedel, J.M., Valcarcel, J. & Lehner, B. Combinatorial Genetics Reveals a Scaling Law for the Effects of Mutations on Splicing. Cell 176, 549–563 e523 (2019).

28. Paronetto, M.P., Passacantilli, I. & Sette, C. Alternative splicing and cell survival: from tissue homeostasis to disease. Cell Death Differ 23, 1919–1929 (2016).

29. Cascino, I., Fiucci, G., Papoff, G. & Ruberti, G. Three functional soluble forms of the human apoptosis-inducing Fas molecule are produced by alternative splicing. J Immunol 154, 2706–2713 (1995).

30. Meynier, S. & Rieux-Laucat, F. FAS and RAS related Apoptosis defects: From autoimmunity to leukemia. Immunol Rev 287, 50–61 (2019).

31. Strasser, A., Jost, P.J. & Nagata, S. The Many Roles of FAS Receptor Signaling in the Immune System. Immunity 30, 180–192 (2009).

32. Sehgal, L., et al. FAS-antisense 1 lncRNA and production of soluble versus membrane Fas in B-cell lymphoma. Leukemia 28, 2376–2387 (2014).

33. Lin, A.Y., Cuttica, M.J., Ison, M.G. & Gordon, L.I. Ibrutinib for chronic lymphocytic leukemia in the setting of respiratory failure from severe COVID-19 infection: Case report and literature review. eJHaem 1, 596–600 (2020).

34. Treon, S.P., et al. The BTK inhibitor ibrutinib may protect against pulmonary injury in COVID-19-infected patients. Blood 135, 1912–1915 (2020).

35. Watanabe-Fukunaga, R., Brannan, C.I., Copeland, N.G., Jenkins, N.A. & Nagata, S. Lymphoproliferation disorder in mice explained by defects in Fas antigen that mediates apoptosis. Nature 356, 314–317 (1992).

36. Fisher, G.H., et al. Dominant interfering Fas gene mutations impair apoptosis in a human autoimmune lymphoproliferative syndrome. Cell 81, 935–946 (1995).

37. Rieux-Laucat, F., et al. Mutations in Fas associated with human lymphoproliferative syndrome and autoimmunity. Science 268, 1347–1349 (1995).

38. Kuehn, H.S., et al. FAS Haploinsufficiency Is a Common Disease Mechanism in the Human Autoimmune Lymphoproliferative Syndrome. The Journal of Immunology 186, 6035 (2011).

39. Price, S., et al. Natural history of autoimmune lymphoproliferative syndrome associated with FAS gene mutations. Blood 123, 1989–1999 (2014).

40. Vuckovic, D., et al. The Polygenic and Monogenic Basis of Blood Traits and Diseases. Cell 182, 1214–1231 e1211 (2020).

41. Berndt, S.I., et al. Genome-wide association study identifies multiple risk loci for chronic lymphocytic leukemia. Nature Genetics 45, 868–876 (2013).

42. Berndt, S.I., et al. Meta-analysis of genome-wide association studies discovers multiple loci for chronic lymphocytic leukemia. Nat Commun 7, 10933 (2016).

43. Hinks, A., et al. Dense genotyping of immune-related disease regions identifies 14 new susceptibility loci for juvenile idiopathic arthritis. Nat Genet 45, 664–669 (2013).

44. Zhou, S., et al. A Neanderthal OAS1 isoform protects individuals of European ancestry against COVID-19 susceptibility and severity. Nature Medicine (2021).

45. Gaziano, L., et al. Actionable druggable genome-wide Mendelian randomization identifies repurposing opportunities for COVID-19. medRxiv, 2020.2011.2019.20234120 (2020).

46. Burgess, S., Thompson, S.G. & Collaboration, C.C.G. Methods for meta-analysis of individual participant data from Mendelian randomisation studies with binary outcomes. Stat Methods Med Res 25, 272–293 (2016).

47. Fortune, M.D., et al. Statistical colocalization of genetic risk variants for related autoimmune diseases in the context of common controls. Nature Genetics 47, 839–846 (2015).

48. Davies, N.M., et al. The many weak instruments problem and Mendelian randomization. Statistics in Medicine 34, 454–468 (2015).

49. Winkler, T.W., et al. Quality control and conduct of genome-wide association meta-analyses. Nature protocols 9, 1192–1212 (2014).

50. Willer, C.J., Li, Y. & Abecasis, G.R. METAL: Fast and efficient meta-analysis of genomewide association scans. Bioinformatics 26, 2190–2191 (2010).

51. Hemani, G., et al. The MR-Base platform supports systematic causal inference across the human phenome. eLife 7, e34408 (2018).

52. Chang, C.C., et al. Second-generation PLINK: rising to the challenge of larger and richer datasets. GigaScience 4, 7–7 (2015).

53. Bowden, J., Davey Smith, G. & Burgess, S. Mendelian randomization with invalid instruments: effect estimation and bias detection through Egger regression. Int J Epidemiol 44, 512–525 (2015).

54. Qingyuan, Z., Jingshu, W., Gibran, H., Jack, B. & Dylan, S.S. Statistical inference in two-sample summary-data Mendelian randomization using robust adjusted profile score. The Annals of Statistics 48, 1742–1769 (2020).

55. Burgess, S., Butterworth, A. & Thompson, S.G. Mendelian Randomization Analysis With Multiple Genetic Variants Using Summarized Data. Genetic Epidemiology 37, 658–665 (2013).

56. Dobin, A., et al. STAR: ultrafast universal RNA-seq aligner. Bioinformatics 29, 15–21 (2013).

57. Li, Y.I., et al. Annotation-free quantification of RNA splicing using LeafCutter. Nature Genetics 50, 151–158 (2018).

58. Taylor-Weiner, A., et al. Scaling computational genomics to millions of individuals with GPUs. Genome Biology 20, 228 (2019).

